# Government messaging about COVID-19 vaccination in Canada and Australia: a Narrative Policy Framework study

**DOI:** 10.1101/2021.12.19.21268042

**Authors:** Freya Saich, Alexandra Martiniuk

## Abstract

**Background:** Storytelling and narratives are critical components to public policy and have been central to public policy communicators throughout the COVID-19 pandemic.

**Aim:** This study applied the Narrative Policy Framework to compare and contrast the policy narratives of the Canadian and Australian Prime Ministers regarding COVID-19 vaccination.

**Methods:** Official media releases, transcripts and speeches published on the websites of Prime Minister Morrison and Prime Minister Trudeau between 31 August 2020 and 10 September 2021 relating to COVID-19 vaccines were thematically analysed according to the Narrative Policy Framework.

**Results:** The policy narratives of Scott Morrison and Justin Trudeau tended towards describing both governments as heroes for securing and rolling out vaccines. Trudeau tended to focus on the villain of COVID-19 while Morrison regularly described other countries as victims of COVID-19 to position Australia as superior in its decision-making. These findings also demonstrate how narratives shifted over time due to changing COVID-19 case numbers, emergence of rare complications associated with the AstraZeneca vaccine and as new information arose.

**Conclusion:** These findings offer lessons for COVID-19 times as well as future pandemics and disease outbreaks by providing insight into how policy narratives influenced policy processes in both Australia and Canada.

## Introduction

Storytelling and narratives are critical components influencing public policy, particularly in a time of crisis.^1-3^ Consequently, policy narratives have been central to public policy communicators throughout the COVID-19 pandemic to convince the public of a problem and legitimise a policy solution.^2,4-8^

With the rapid development and rollout of COVID-19 vaccines, policy actors have been required to deploy narratives to convince the public of their safety and efficacy, and encourage the population to get vaccinated. However, significant media and expert attention has been drawn to COVID-19 vaccine hesitancy.^9^ Studies regarding the uptake of COVID-19 vaccines have largely focused on individuals’ views, their trust in governments, and the information they receive.^10^ There has been limited critical exploration of government policy and the narratives that have been deployed surrounding COVID-19 vaccines, particularly between countries with differing epidemiological profiles. This is a critical area of study as facts, evidence and data often do little in isolation to convince the public.^1^ Instead, it is typically the narrative used to weave together the context, problem and resolution, combined with evidence, that influences opinions and behaviours.^1^

The Narrative Policy Framework is a relatively new framework that applies scientific techniques to the study of policy narratives.^1^ The Narrative Policy Framework is premised on the fact that narratives must have distinct components including settings (bounded by time, geography or other characteristics), characters (hero, villain and victims that may either be a human, place or thing), plots (which connect the characters to the setting) and a moral of the story (policy solution).^1,11^ This framework has been used to analyse the influence of policy narratives on individual decision-making (micro), their effect on the policy processes (meso), and how policy narratives amongst cultures and institutions change over time (macro).^1,11^

To date, most Narrative Policy Framework studies conducted in the context of COVID-19 have focused on policy narratives at the onset of the pandemic and no studies have applied this framework to policy narratives in relation to COVID-19 vaccines.^4,7-8^ This is worth further exploration as the dynamic nature of the pandemic has meant that the narratives used by governments have also had to shift with the advent of new variants, research and medical developments and as new information arises.

The Narrative Policy Framework requires a deep understanding of the cultural setting in which the narrative is constructed.^1,11^ Canada and Australia share many similarities; both are federated nations comprised of provinces/states and territories, and both Federal Governments have been responsible for COVID-19 vaccine procurement, approvals and providing scientific advice, while provincial, state and territorial governments have been largely responsible for administering the vaccines.^12-13^ Politically, Australia and Canada differ in their leadership at the federal level with Prime Minister Justin Trudeau (Canada) the leader of the left-leaning Liberal Party whereas Prime Minister Scott Morrison (Australia) is the leader of the right-leaning Liberal Party.^14-15^

The COVID-19 experience has been vastly different in Canada and Australia. As of 1 November 2021, Canada recorded a total of 1.7 million COVID-19 cases and approximately 29,000 deaths in a population of 38.2 million.^16^ In comparison, Australia has been relatively successful on a global scale at controlling COVID-19, recording 170,000 cases and over 1,700 deaths within the same period in a population of 25.9 million.^17^

### Aim

This study applies the Narrative Policy Framework to compare and contrast narratives deployed by both the Canadian and Australian Prime Ministers regarding COVID-19 vaccines to convince the public of their safety and efficacy and encourage citizens to get vaccinated.

### Methodology

Official media releases, speeches and press conferences published on the websites of Prime Minister Morrison^18^ and Prime Minister Trudeau^19^ published between 31 August 2020 and 10 September 2021 were reviewed. To be included for analysis, materials were required to reference COVID-19 vaccines or vaccinations. In accordance with the Narrative Policy Framework these materials were also required to include at least one character (hero, villain or victim) and a plot to be included for analysis.^11^ As this study was focused specifically on vaccination communication and uptake in Australia and Canada, materials that only focused on donation of COVID-19 vaccines or global meetings of leaders discussing COVID-19 vaccines were excluded. If a document contained excerpts from other Ministers or Health Officials, that material was excluded and only the excerpts from Morrison or Trudeau were included for content analysis.

In total 216 materials met the selection criteria (87 for Canada and 129 for Australia). A qualitative methodology for analysing these data was chosen to explore the nuances and framing of the narratives. These materials were organised according to phase: procurement, approval, early rollout, AstraZeneca blood clots and scale-up of rollout. Within these phases, Microsoft Excel was used to code the differing characters between Canada and Australia as well as the overall plots and moral of the story (policy solutions) as described by Trudeau and Morrison.

## Results

### Vaccine procurement

In this phase, Trudeau’s plot embodied that of ‘stymied progress’ – that is to say that the first wave of COVID-19 cases was terrible, but then the situation improved due to a hero (Federal Government) but COVID-19 was worsening again emphasising the need for the Federal Government to procure vaccines.^11^ By 23 October 2020, eight vaccine agreements were signed securing a total of 358 million doses resulting in “the most diverse vaccine portfolio of any country”.^20^ Canada also pursued sovereign vaccine manufacturing and capability, as part of the “made-in-Canada” approach to the pandemic.^21^ Despite this, Trudeau reminded Canadians on four occasions that a “vaccine won’t help you – or your family – if you get COVID-19 now” encouraging Canadians to adhere to the public health restrictions to protect others and save lives.^21^

Building on Weible and Schalger’s^22^ plot descriptions, the Australian phase of vaccine procurement will be described as ‘near complete victory’ as the villain (COVID-19) is never truly vanquished and thus vaccines were needed to maintain this victory which is achieved by the hero (Morrison and his government). To illustrate this, between 19 August and 5 November 2020 Morrison announced four vaccine deals. Similar to Canada, Morrison couched two of the vaccine deals (AstraZeneca and University of Queensland) as part of a “home-grown sovereign vaccine plan”^23^ that would utilise local manufacturing that is amongst the “best in the world” in order to secure supply chains.^24^ Morrison said this placed Australia amongst an “elite circle of countries that has been able to respond on behalf of their population” with just 20 countries able to manufacture COVID-19 vaccines.^25^

These announcements were largely devoid of explicit mention of a villain and instead emphasised the heroic nature of Australia having heeded the medical advice of experts and controlling COVID-19 infections earning it a ranking among the leading economies in the world. The vaccine announcements sought to further this leading position to “keep Australia right up the front”, without taking any shortcuts on safety.^23^

### Regulatory approval

The stymied progress plot continued into this phase for Canada. On 9 December 2020, the Pfizer vaccine was the first vaccine to be approved in Canada. This came in the midst of Canada’s second wave – at the beginning of winter in the Northern Hemisphere – when cases and hospitalisations were the highest in the pandemic thus far. As a result, the development and approval of the vaccines was regularly described by Trudeau as the “light at the end of the tunnel” and as a symbol of hope to Canadians, thus Trudeau’s government was the hero for securing these vaccines.^26^ In the lead up to the approval, Trudeau emphasised “Canada’s globally recognized gold standard for medical approvals” and that “any vaccine approved in Canada will be safe and effective”.^27^ As subsequent vaccines were approved in December 2020, February 2021 and March 2021, Trudeau again stated “the best experts have upheld this highest standard”^28^ and that they have each been “independently approved”.^29^ The moral was thus implicit that Canadians should be confident to get vaccinated.

It was not until the 25 January 2021 that the first vaccine, Pfizer, was approved in Australia with the AstraZeneca vaccine approved nearly a month later on 16 February 2021. On 12 occasions, Morrison stated that neither of these vaccines had been approved under an emergency approval process, as had occurred in many European nations such as the UK. Morrison used a ‘blame the victim’ plot^11^ to describe how COVID-19 was out of control in these countries and were thus victims with no choice but to ‘cut corners’ on regulatory approvals by rushing them through emergency approval processes. Morrison also maintained the ‘near complete victory’ plot by positioning Australia in stark contrast by not taking any “short cuts” on the approval process.^30^ Australia was positioned as a hero because of “our amazing efforts”^31^ and our “2020 achievements”^32^ where the virus was successfully suppressed and controlled. Australia was a hero as “we are one of only a handful of countries to have gone through such a comprehensive and thorough level of oversight to ensure the vaccines are safe”^30^ having been “overseen by the best medical experts in the world” at the TGA.^33^ Morrison regularly stated that Australians could have confidence in the vaccine and that no one was being put at risk (moral of the story).

Months later in August, Moderna was approved. At this point the narrative shifted from not rushing approvals, to the TGA expediting approvals by “accepting rolling data submissions for review, collaboration with international regulators, and proactive work with the sponsor, Moderna.”^34^

### Beginning of rollout

Ahead of the rollout Trudeau emphasised that as soon as the doses were approved, Canada would be ready. In line with these statements just a few days after Pfizer was approved in Canada, doses arrived in the country and the rollout began. Throughout this phase, Trudeau regularly described the Federal Government as the hero for working “tirelessly”^35^ and “around the clock”.^36^ On 14 occasions, during this phase, Trudeau stated that the Federal Government was working with the provinces and territories to get the vaccine delivered, distributed and administered ‘as quickly as possible’ and noted this as an example of a true ‘Team Canada’ approach. He also stated that the “Federal Government is covering the cost of the vaccines and the supplies needed to administer them” again emphasising the heroic nature of the Federal Government in securing and paying for the vaccines.^37^ Throughout this time, COVID-19, the pandemic, ‘the virus’ and variants – framed as the villain – were used to encourage Canadians to get vaccinated while employing other measures (distancing, mask wearing, handwashing etc.) to get through the ‘long winter’. While there were some delays with the Pfizer doses, Trudeau described these as “temporary, short-term issues” that were “to be expected”^38^ and despite this, things were “going as expected”.^39^

Nearly a month after receiving approval from the TGA, the first Pfizer doses arrived in Australia on 15 February 2021. Morrison sought to allay concerns about the vaccines, stressing the “care that is being provided to every Australian through the amazing processes and systems that have been built here”.^40^ At the same time Morrison stated, “these are all great vaccines, they’re not better than each other”.^40^ Morrison continued the ‘near complete victory’ plot by describing how the “Australian way through this pandemic”^41^ had led to greater success than other countries and called on Australians to “continue on the successful path we’ve been on” by getting vaccinated (moral of the story).^42^ Morrison also maintained the ‘blame the victim’ plot when Italy blocked supply of Pfizer doses to Australia, describing how Italy “are in an unbridled crisis situation”.^43^ This furthered Morrison’s argument for sovereign vaccine production of the AstraZeneca vaccine, and on 12 occasions Morrison reiterated that few countries had that capability, making Australia “part of a small club”.^44^ Furthermore, Morrison stated that “were it not for the fact that our Government took the strategic decision last August to not rely on international supply of vaccines, then Australia would not have a vaccination programme now.”^45^

As the rollout continued and as criticism mounted that Australia was not meeting its early vaccination targets, Morrison reverted to emphasising safety stating, “It’s not a race. It’s not a competition…”^46^ and “I don’t intend to rush the proces”^47^ and detailed the processes that needed to be followed by the TGA before administration to the population. By April, Morrison began comparing Australia’s rollout to comparable countries such as Germany, Canada, Japan, South Korea and New Zealand, stating that Australia was either on par or further advanced than they had been at the same stage in their rollout and “most of the world would want to be right here at the moment” given the lack of community transmission and relative freedoms across the country.^48^

### Health advice on the use of the AstraZeneca vaccine

There was no explicit mention by Trudeau in any of the reviewed documents of thrombosis with thrombocytopenia (blood clots) associated with the AstraZeneca vaccine. Instead on 19 March 2021, Trudeau reiterated that any vaccine approved in Canada, by Health Canada, was safe and effective and that Canadians “can feel very confident getting it” and confirmed to Canadians that “the best vaccine for you is the first one you’re offered”.^49^ Following this, Trudeau stated that he and his wife had both received AstraZeneca and were “feeling great. We’re feeling more protected, and we’re also feeling like we’re part of the solution going forward.”^50^

In comparison, Scott Morrison held a late-night press conference on 8 April 2021 to announce that the Australian Technical Advisory Group on Immunisation (ATAGI) had advised a preference to vaccinate people under 50 with Pfizer (thus over 50s would eligible only for AstraZeneca). The next day, Morrison confirmed this was not “a ban on the AstraZeneca vaccine. It is not a prohibition on the AstraZeneca vaccine” and said that his mother would be vaccinated with AstraZeneca in a couple of weeks.^51^ Given that less than four percent of the population had received a COVID-19 vaccine and the rollout had largely been contingent on the use of AstraZeneca, the program would need to undergo “recalibration”.^51^ Nonetheless Morrison emphasised how the AstraZeneca vaccine was appropriate for people over 50, of which people over 70 were most vulnerable (and thus victims) as evidenced by Victoria’s second wave in 2020. On 17 June the ATAGI advice changed again preferring Pfizer for people under 60. On 28 June 2021 Morrison made an unexpected announcement stating that anyone (regardless of age) could discuss the AstraZeneca vaccine with their doctor and receive it with informed consent. It was only on 13 July that ATAGI officially made this recommendation.

The Prime Minister conceded several times that the changing advice regarding this AstraZeneca vaccine “caused some hesitation”^52^ and that Australia was “prisoners of our own success”.^53^ To encourage uptake of the AstraZeneca vaccine, Morrison noted on several occasions that “we saw it save countless number of lives in the United Kingdom, and it will save lives here, too”,^54^ a fundamental shift from the ‘blame the victim’ plot having previously described the UK as helpless in comparison to Australia.

### Scale-up of roll-out

On 19 February 2021, Trudeau declared that Canada was in the ramp-up phase of the vaccination program. With substantial increases in supply and as mass vaccinations commenced, “better days are ahead”.^55^ Trudeau encouraged Canadians who had already been vaccinated “to keep doing their part to keep others safe as we continue to increase vaccinations”.^56^ As of 4 May 2021, COVID-19 case numbers were falling quickly. Trudeau used this to illustrate the effectiveness of the vaccines: “we’re seeing how vaccines – along with public health measures – keep people safe”.^57^

By 25 May, Canada had reached a ‘turning point’ in its vaccination rollout with over half of the population having received at least one dose. At this point, the narrative of Canada as hero reemerged after being ranked third amongst G20 countries for total proportion of the population vaccinated. By 4 June Canada was first amongst the G20 nations and even overtook the US on 19 July. Trudeau again described these achievements as a “true Team Canada approach”.^58^ The promise of a “better summer” sought to encourage Canadians to continue to get vaccinated and get back to the things they had previously enjoyed.^59^ Despite transmission continuing to occur, Trudeau applied the ‘complete victory’ plot stating that “Canada has come through this pandemic better than almost any other country in the world. And it’s because we’ve been there for each other” adding weight to Canada’s collaborative narrative to increasing vaccinations.^60^

From July 2021, Morrison’s narrative experienced a fundamental shift with outbreaks of the Delta variant on Australia’s east coast in New South Wales, Victoria and the Australian Capital Territory that resulted in lockdowns that would last until October 2021. Similar to Trudeau’s comments over the 2020-21 winter, vaccines were presented as one strategy to suppress COVID-19 infections with the tools of contact tracing and testing no longer proving sufficient to control COVID-19. Vaccines were also described as the “light at the end of the tunnel” and “doses of hope”, similar to how Trudeau described them in 2020-21.^61-63^

In August 2021, with one third of the population having received one dose of a COVID-19 vaccine, Morrison acknowledged that there had been some setbacks in the rollout but that it was also his responsibility to fix these problems. Morrison was again a hero for leaving “no stone was left unturned in the global search to find additional vaccines” having secured deals with foreign governments that led to substantial increases in supply.^63^ By 19 August 2021, Morrison declared that Australia had reached a ‘turning point’ in its vaccination rollout – reaffirming the ‘near complete victory’ plot – and by late August detailed how Australia was again a hero for exceeding the peak of the UK and US vaccination programs and as a result had “done better than almost any other country in the world in saving lives and livelihoods.”^64^

## Discussion and implications

This study has offered a unique use of the Narrative Policy Framework to explore the narratives used to convince the public to get vaccinated against COVID-19 and has added to the growing application of this framework in times of crisis and specifically to the COVID-19 pandemic.^3,8^ This study has enabled an in-depth exploration of the various characters, plots and policy solutions used by Morrison and Trudeau in convincing the public of the safety and efficacy of the vaccines, and the importance of vaccines and getting vaccinated.

Both Morrison and Trudeau tended towards describing their governments as heroes for ensuring vaccinations were procured, approved and rolled out across the country whereas the resounding villain was the ‘the virus’, COVID-19 disease and variants. Various populations at risk of severe disease or death associated with COVID-19 were positioned as ‘victims’ and in need of protection either through the direct use of vaccines or by those around them. Morrison also described whole countries (particularly the UK) as victims for their poor policy decisions and inability to control COVID-19 to legitimise Australia’s policy decisions about the vaccine approvals and to defend delays in the early stages of the rollout. In doing so, Morrison created a paradox whereby these same countries were later slated as leaders to be emulated. For instance, Morrison used the UK’s vaccination rate in mid-2021 as a goal to achieve for Australia’s vaccination program in the scale-up phase of the rollout and to promote the use of the AstraZeneca vaccine, given its widespread use in the UK. This highlights the dynamic nature of policy narratives and the strategic selecting in and out of characters to advance the policy narrative at the time.^65^

In terms of plots, Trudeau’s tended to utilise that of ‘stymied progress’ with peaks and troughs of case numbers, deaths and hospitalisations and measures needed to bring COVID-19 under control. However, in the ramp-up phase the plot shifted to reflect ‘complete victory’ over the virus through Canada’s collaborative approach to vaccinating the nation.

With the exception of the Delta outbreak and ensuing wave in July to October, Morrison’s narrative used a plot to centre discussions around the ‘near complete victory’ of COVID-19 in Australia. However, this made the policy solution (to get vaccinated) somewhat unclear as Australians were encouraged to trust in the process and the vaccines, although many were ineligible or confused by the changing official advice as well as advice offered by Morrison. This highlights the importance of coherent narratives as well as narratives being matched with the appropriate resources for them to succeed.^4^

Canada also received criticism for its handling of the AstraZeneca vaccine. While it was unable to be explored in this study given the focus on the national policy narrative, some of the provincial governments suspended the use of the AstraZeneca vaccine for first doses.^66^ Furthermore, the Canadian National Advisory Committee on Immunization (NACI) had recommended that mRNA vaccines were preferred over viral-vector vaccines such as AstraZeneca, despite Trudeau continuing to emphasise that all vaccine types were safe and effective.^67^

Both countries’ handling of their advice around the AstraZeneca vaccine offers a cautionary tale for future risk communication given the confusion caused by rapidly changing advice. It is also worth considering the influence of the Australian narrative as a hero and having achieved ‘near complete control’ of COVID-19 and whether this led to a lack of urgency and complacency within both policy decisions and amongst individuals.

Given that policy narratives are essential to policy processes and decisions,^68^ these findings also demonstrate the inherent complexity of making ‘policy on the run’. From the outset Canada chose to procure as many vaccine candidates as possible. While this has been met with criticism for being greedy with the highest number of vaccines per capita,^69^ it nonetheless protected Canada from

Australia’s situation in relying ultimately on just two vaccines (for the most part of 2021) and the issues that ensued with supply, rare medical complications that did not arise until mass vaccination and a changing risk/benefit profile of vaccine types either due to rare adverse effects and/or COVID-19 variants.

It is also clear how the ‘near complete victory’ narrative influenced decision-making particularly in Australia in not seeking to ‘rush’ the rollout initially, but then by May 2021 Australia was prepared to expedite the approval process of the Moderna vaccine. It is worth comparing this to another country that has fared similar to Australia, Singapore, which commenced its vaccination program at the end of December 2020 when case numbers were low and was able to vaccinate upwards to 80% of the total population while simultaneously keeping case numbers low.^70^ While this warrants further attention, it nonetheless adds further evidence to the role of narratives in policy processes and decision-making and subsequent realities on the ground.

## Limitations

There are several limitations of this study. The study design did not include the narratives used by other Federal Ministers or State, Provincial and Territory leaders or officials and how these narratives may echo or diverge from that of Morrison and Trudeau. This may be particularly important in the Australian context whereby the pandemic has resulted in greater trust in state and territory leaders and declining trust in the Federal Government.^71^ In Canada, trust in both the Prime Minister and provincial premiers has declined which may be reflective of the lack of success in controlling COVID-19 in Canada compared to Australia.^72^ Future research could assess the effectiveness of similar policy narratives in influencing public opinion and intention to vaccinate. Lastly, we are unable to conclude whether one approach or another policy narrative approach was more successful using, for instance, overall vaccination rates by time, given the likely confounding role of changing medical advice, supply issues, cultural and COVID-19 related factors.

## Conclusion

This study has explored the differing narratives used by Trudeau and Morrison to encourage vaccination uptake in the context of divergent epidemiological profiles in the COVID-19 pandemic. This adds to the growing use of the Narrative Policy Framework to analyse and understand policy narrative during crises and uniquely in regards to COVID-19 vaccines. While this study highlights areas for further research, these findings offer lessons for future pandemics and disease outbreaks particularly from the examples of conflicting plots and character portrayals in the policy narratives of both Australia and Canada which likely influenced stakeholders and the general public’s decision-making and actions.

## Data Availability

All data produced are available online and links are contained within the manuscript.

https://pm.gc.ca/en/news

https://www.pm.gov.au/media

## References

[1] Jones, M.D., McBeth, M.K. and Shanahan, E.A. (eds) (2014) ‘Introducing the Narrative Policy Framework’ in The Science of Stories: Applications of the Narrative Policy Framework in Public Policy Analysis, Palgrave Macmillan.

[2] Montiel, C.J., Uyheng, J. and Dela Paz, E. (2021) The Language of Pandemic Leaderships: Mapping Political Rhetoric During the COVID-19 Outbreak, Political Psychology, 42:747–766.

[3] Crow, D., Lawhon, L., Berggren, J., Huda, J., Koebele, E. and Kroepsch, A. (2017) A Narrative Policy Framework Analysis of Wildfire Policy Discussions in Two Colorado Communities, Politics and Policy, 45(4):626–656.

[4] Mintrom, M. and O’Connor (2020) The importance of policy narrative: effective government responses to Covid-19, Policy Design and Practice, 3(3):205–227.

[5] Kuhlmann, J. and Blum, S. (2021) Narrative plots for regulatory, distributive, and redistributive policies, European Policy Analysis, 7(S):276–302.

[6] O’Donovan, K. (2018) Does the Narrative Policy Framework Apply to Local Policy Issues?, Politics and Policy, 46(4),:532–570.

[7] Mintrom, M., Rost Rublee, M., Bonotti, M. and Zech, S. (2021) Policy narratives, localisation, and public justification: responses to COVID-19, Journal of European Public Policy, 28:1219–1237.

[8] Apriliyanti, I.D., Utomo, V. P. and Purwanto, E.A. (2021) Examining the policy narratives and the role of the media in policy responses to the COVID-19 crisis in Indonesia, Journal of Asian Public Policy, online.

[9] Rosenbaum, L. (2021) Escaping Catch-22 — Overcoming Covid Vaccine Hesitancy, The New England Journal of Medicine, 384(14):1367–1371.

[10] Richardson, L..M and Thaker, J.T. (2021), Australia’s COVID-19 Vaccine Audiences, Monash University & Massey University, Melbourne, Australia.

[11] Shanahan, E., Jones, M.D., McBeth, M. (2018) How to conduct a Narrative Policy Framework study, The Social Science Journal, 55:332–345.

[12] Government of Canada (2021) COVID-19 immunization: Federal, provincial and territorial statement of common principles. Available from: https://www.canada.ca/en/public-health/services/diseases/2019-novel-coronavirus-infection/canadas-reponse/covid-19-immunization-federal-provincial-territorial-statement-common-principles.html accessed 3 November 2021.

[13] Australian Government (2020) Australian COVID-19 Vaccination Policy. Available from: https://www.health.gov.au/sites/default/files/documents/2020/12/covid-19-vaccination-australian-covid-19-vaccination-policy.pdf, accessed 2 November 2021.

[14] Wallenfeldt, J. (2021) Justin Trudeau, Britannica. Available from: https://www.britannica.com/biography/Justin-Trudeau, accessed 4 November 2021.

[15] Liberal Party (2021) Our beliefs. Available from: https://www.liberal.org.au/our-beliefs, accessed 4 November 2021.

[16] Government of Canada (2021) COVID-19 daily epidemiology update. Available from: https://health-infobase.canada.ca/covid-19/epidemiological-summary-covid-19-cases.html accessed 2 November 2021.

[17] Department of Health (2021) COVID-19 at a glance, Australian Government. Available from: https://www.health.gov.au/sites/default/files/documents/2021/11/coronavirus-covid-19-at-a-glance-1-november-2021.pdf accessed 2 November 2021.

[18] Prime Minister of Australia (2021), Media Centre. Available from: https://www.pm.gov.au/media

[19] Trudeau, J. (2021) News. Available from: https://pm.gc.ca/en/news

[20] Trudeau, J. (2020) Statement by the Prime Minister to mark the New Year, Statement. Available from: https://pm.gc.ca/en/news/statements/2020/12/31/statement-prime-minister-mark-new-year

[21] Trudeau, J. (2020) Prime Minister’s remarks on the COVID-19 situation and support for Veterans, Speech, Available from: https://pm.gc.ca/en/news/speeches/2020/11/10/prime-ministers-remarks-covid-19-situation-and-support-veterans

[22] Weible, C.M. and Schalger, E. (2014) ‘Narrative Policy Framework: Contribution, Limitations and Recommendations’ in Jones, M.D., McBeth, M.K. and Shanahan, E.A. (eds) The Science of Stories: Applications of the Narrative Policy Framework in Public Policy Analysis, Palgrave Macmillan.

[23] Prime Minister (2020) Press Conference - Australian Parliament House, ACT, Press Conference. Available from: https://www.pm.gov.au/media/press-conference-australian-parliament-house-act-070920

[24] Prime Minster, Minister for Health, Minister for Industry, Science and Technology (2020), New deal secures potential COVID-19 vaccine for every Australian, Media Release. Available from: https://www.pm.gov.au/media/new-deal-secures-potential-covid-19-vaccine-every-australian

[25] Prime Minister (2020) Press Conference - Australian Parliament House, ACT, Press Conference. Available from: https://www.pm.gov.au/media/press-conference-australian-parliament-house-act-32

[26] Trudeau, J. (2020) Prime Minister’s remarks updating Canadians on the COVID-19 situation and announcing new supports for Indigenous communities, Speech. Available from: https://pm.gc.ca/en/news/speeches/2020/11/27/prime-ministers-remarks-updating-canadians-covid-19-situation-and

[27] Trudeau, J. (2020) Prime Minister’s remarks announcing the early delivery of the Pfizer-BioNTech COVID-19 vaccine, Speech. Available from: https://pm.gc.ca/en/news/speeches/2020/12/07/prime-ministers-remarks-announcing-early-delivery-pfizer-biontech-covid-19

[28] Trudeau, J. (2020) Prime Minister’s remarks updating Canadians on COVID-19 and announcing the upcoming appointment of the new Chief of the Defence Staff, Speech. Available from: https://pm.gc.ca/en/news/speeches/2020/12/23/prime-ministers-remarks-updating-canadians-covid-19-and-announcing

[29] Trudeau, J. (2020) Prime Minister’s remarks updating Canadians on COVID-19 vaccine authorizations and procurement, Speech. Available from: https://pm.gc.ca/en/news/speeches/2021/02/26/prime-ministers-remarks-updating-canadians-covid-19-vaccine-authorizations

[30] Prime Minister (2021) Press Conference. Available from: https://www.pm.gov.au/media/press-conference-2

[31] Prime Minister (2020) Doorstop - Riverside, TAS. Available from: https://www.pm.gov.au/media/doorstop-riverside-tas

[32] Prime Minister (2021) Address - National Press Club Barton ACT. Available from: https://www.pm.gov.au/media/address-national-press-club-barton-act

[33] Prime Minister (2021) Doorstop-Symonston, ACT, Media Release. Available from: https://www.pm.gov.au/media/doorstop-symonston-act

[34] Prime Minister, Minister for Health and Aged Care (2021) Moderna COVID-19 vaccine approved for use in Australia, Media Release. Available from: https://www.pm.gov.au/media/moderna-covid-19-vaccine-approved-use-australia

[35] Trudeau, J. (2021) Prime Minister’s remarks updating Canadians on the COVID-19 situation and support for Canadian businesses, Speech. Available from: https://pm.gc.ca/en/news/speeches/2021/01/26/prime-ministers-remarks-updating-canadians-covid-19-situation-and-support

[36] Trudeau, J. (2021) Prime Minister’s remarks updating Canadians on the COVID-19 situation and vaccine rollout, Speech. Available from: https://pm.gc.ca/en/news/speeches/2021/02/05/prime-ministers-remarks-updating-canadians-covid-19-situation-and-vaccine

[37] Trudeau, J. (2021) Prime Minister’s remarks updating Canadians on the COVID-19 situation, vaccines, and travel restrictions, Speech. Available from: https://pm.gc.ca/en/news/speeches/2021/01/05/prime-ministers-remarks-updating-canadians-covid-19-situation-vaccines-and

[38] Trudeau, J. (2021) Prime Minister’s remarks updating Canadians on the COVID-19 situation and on tax filing, Speech. Available from: https://pm.gc.ca/en/news/speeches/2021/02/09/prime-ministers-remarks-updating-canadians-covid-19-situation-and-tax

[39] Trudeau, J. (2021) Prime Minister’s remarks updating Canadians on COVID-19, vaccines, and affordable housing, Speech. Available from: https://pm.gc.ca/en/news/speeches/2021/01/15/prime-ministers-remarks-updating-canadians-covid-19-vaccines-and

[40] Prime Minister (2021) Press Conference – Camperdown NSW. Available from: https://www.pm.gov.au/media/press-conference-camperdown-nsw

[41] Prime Minister, Minister for Health and Aged Care (2021) First COVID-19 Vaccinations, Media Release, Available from: https://www.pm.gov.au/media/first-covid-19-vaccinations

[42] Prime Minister (2021) Press Conference – Castle Hill, NSW, Media Release. Available from: https://www.pm.gov.au/media/press-conference-castle-hill-nsw

[43] Prime Minister (2021) Press Conference – Sydney, NSW, Transcript. Available from: https://www.pm.gov.au/media/press-conference-sydney-nsw-7

[44] Prime Minister (2021) Doorstop – Parkville, VIC, Transcript. Available from: https://www.pm.gov.au/media/doorstop-parkville-vic

[45] Prime Minister (2021) Press Conference – Australian Parliament House, ACT, Transcript. Available from: https://www.pm.gov.au/media/press-conference-australian-parliament-house-act-36

[46] Prime Minister (2021) Doorstop – Sydney Domestic Airport, NSW, Media Release. Available from: https://www.pm.gov.au/media/doorstop-sydney-domestic-airport-nsw

[47] Prime Minister (2021) Press Conference – Australian Parliament House, ACT, Transcript. Available from: https://www.pm.gov.au/media/press-conference-australian-parliament-house-act-060421

[48] Prime Minister (2021) Press Conference – Australian Parliament House, ACT, Transcript. Available from: https://www.pm.gov.au/media/press-conference-australian-parliament-house-act-070421

[49] Trudeau, J. (2021) Prime Minister’s remarks on COVID-19 vaccines and affordable housing investments, Speech. Available from: https://pm.gc.ca/en/news/speeches/2021/03/19/prime-ministers-remarks-covid-19-vaccines-and-affordable-housing

[50] Trudeau, J. (2021) Prime Minister’s remarks on the COVID-19 situation and the Budget 2021 plan for a feminist recovery, Speech. Available from: https://pm.gc.ca/en/news/speeches/2021/04/27/prime-ministers-remarks-covid-19-situation-and-budget-2021-plan-feminist

[51] Prime Minister (2021) Press Conference – Australian Parliament House, ACT, Transcript. Available from: https://www.pm.gov.au/media/press-conference-australian-parliament-house-act-09april21

[52] Prime Minister (2021) Press Conference – Canberra, ACT, Transcript. Available from: https://www.pm.gov.au/media/press-conference-canberra-act-6

[53] Prime Minister (2021) Transcript - Press Conference - Australian Parliament House, ACT. Available from: https://www.pm.gov.au/media/transcript-press-conference-australian-parliament-house-act

[54] Prime Minister (2021) Press Conference, Transcript. Available from: https://www.pm.gov.au/media/press-conference-6

[55] Trudeau, J. (2021), First Ministers mark the first anniversary of the global COVID-19 pandemic, Media Release. Available from: https://pm.gc.ca/en/news/news-releases/2021/03/11/first-ministers-mark-first-anniversary-global-covid-19-pandemic

[56] Trudeau, J. (2021) Prime Minister’s remarks on COVID-19 vaccines, the lifting of long-term drinking water advisories, and support for Canadians, Speech. Available from: https://pm.gc.ca/en/news/speeches/2021/03/12/prime-ministers-remarks-covid-19-vaccines-lifting-long-term-drinking-water

[57] Trudeau, J. (2021) Prime Minister’s remarks on the COVID-19 situation and Budget 2021, Speech. Available from: https://pm.gc.ca/en/news/speeches/2021/05/04/prime-ministers-remarks-covid-19-situation-and-budget-2021

[58] Trudeau, J. (2021) Canada reaches major vaccine campaign milestone, Media Release. Available from: https://pm.gc.ca/en/news/news-releases/2021/07/27/canada-reaches-major-vaccine-campaign-milestone

[59] Trudeau, J. (2021) Prime Minister’s remarks updating Canadians on the COVID-19 situation and support for Manitoba, Speech. Available from: https://pm.gc.ca/en/news/speeches/2021/05/25/prime-ministers-remarks-updating-canadians-covid-19-situation-and-support

[60] Trudeau, J. (2021) Prime Minister’s remarks announcing major vaccine campaign milestone for Canada, Speech. Available from: https://pm.gc.ca/en/news/speeches/2021/07/27/prime-ministers-remarks-announcing-major-vaccine-campaign-milestone-canada

[61] Prime Minister (2021) Time to Shift Focus from Case Numbers to Hospitalisations, Opinion. Available from: https://www.pm.gov.au/media/time-shift-focus-case-numbers-hospitalisations

[62] Prime Minister (2021) Press Conference – Canberra, ACT, Transcript. Available from: https://www.pm.gov.au/media/press-conference-canberra-act-150821

[63] Prime Minister, Minister for Foreign Affairs, Minister for Health and Aged Care (2021) Four million Pfizer doses arrive to super-charge vaccine roll out. Available from: https://www.pm.gov.au/media/four-million-pfizer-doses-arrive-super-charge-vaccine-roll-out

[64] Prime Minister (2021) Vaccination Targets - 2 August 2021, Opinion. Available from: https://www.pm.gov.au/media/vaccination-targets-2-august-2021

[65] Lowndes, V. (2016) Storytelling and narrative in policymaking, in Stoker, G. and Evans, M. (eds), Evidence Based Policymaking in the Social Sciences: Methods that Matter. Policy Press, Bristol, p.103.

[66] Tasker, J.P. (2021) As some provinces halt the use of AstraZeneca, Canada confirms 655,000 more doses will arrive next week, CBC. Available from: https://www.cbc.ca/news/politics/astrazeneca-shots-delivery-1.6024063, accessed 4 November 2021.

[67] Jackson, S. (2021) NACI advice to wait for ‘preferred vaccine’ sends bad message to essential workers: doctor, CBC. Available from: https://www.cbc.ca/radio/asithappens/as-it-happens-tuesday-edition-1.6013354/naci-advice-to-wait-for-preferred-vaccine-sends-bad-message-to-essential-workers-doctor-1.6013526 accessed 4 November 2021.

[68] Shanahan, E., Jones, M. and McBeth, M. (2011) Policy Narratives and Policy Processes, Policy Studies Journal, 39(3):535–561.

[69] Houston, A.R. and Murphy, S. (2021) Canada is no global health leader on COVID-19 vaccine equity, The Lancet, 397:1803.

[70] Aravindan, A. (2020) Singapore begins rollout of Pfizer’s COVID-19 vaccine with healthcare workers, Reuters. Available from: https://www.reuters.com/business/healthcare-pharmaceuticals/singapore-begins-rollout-pfizers-covid-19-vaccine-with-healthcare-workers-2020-12-30/, accessed 4 November 2021.

[71] Browne, B. (2021) State revival: The role of the states in Australia’s COVID-19 response and beyond, The Australia Institute. Available from: https://australiainstitute.org.au/wp-content/uploads/2021/07/P1055-State-revival-WEB.pdf, accessed 2 November 2021.

[72] Tumilty, R. (2021) COVID pandemic corroded Canadians’ trust in politicians — even their neighbours, poll finds, National Post. Available from: https://nationalpost.com/news/politics/covid-pandemic-eroded-canadians-trust-in-politicians-science-and-even-their-neighbours-poll-finds accessed 4 November 2021.

